# Speeding and Traffic-Related Injuries and Fatalities during the 2020 COVID-19 Pandemic: The Cases of Seattle and New York City

**DOI:** 10.1101/2021.08.08.21261745

**Authors:** Felix Haifeng Liao, Michael Lowry

## Abstract

Despite fewer cars on roads during the COVID-19 pandemic, deaths associated with motor vehicle collisions in New York City and Seattle remained largely unchanged in 2020. Using police data on weekly counts of collisions, we compared trends in 2020 with those of 2019, while controlling for the reduction of traffic volumes and seasonal weather conditions. Results of difference-in-differences estimation suggest that during the early months of the pandemic, or March-May, the incidence rates of severe or fatal injury crashes related to speeding increased by nearly 8 times in Seattle and more than 4 times in New York City. In the rest of 2020, they were still significantly higher than what would be expected in the absence of the pandemic. This research suggests that in similar situations that depress travel demand (e.g., another pandemic), policymakers should formulate plans to reduce speeding which may prevent an upswing in severe injuries and fatalities.

## BACKGROUND

As of July 2021, the outbreak of COVID-19 has resulted in over 4 million deaths globally and affects many aspects of human life. Other health outcomes as a result of the crisis, such as increases in suicides and drug overdoses, have also been reported.^1,2^ During the early months of the pandemic, public health responses, notably stay-at-home orders, were implemented to contain the spread of the virus. These control measures resulted in a substantial reduction in traffic volume and a simultaneous decrease in motor vehicle collisions (MVCs).^3–8^ The decrease in total crashes, however, does not necessarily result in fewer fatalities on roads. A study found that there was a surge of fatal single-vehicle crashes in Connecticut during March-April of 2020 when the lockdown order was issued.^9^ Evidence from Japan suggested that drivers were more likely to commit speeding-related violations that caused fatal crashes in the second month of the lockdown.^10^ A recent report from the U.S. Federal Highway Administration indicated that although all miles driven on roads and highways reached its lowest point in the last two decades, the deaths associated with MVCs in the U.S. increased by 8% in 2020.^11^

Drawing upon the cases of Seattle and New York City (NYC), the two U.S. cities impacted early by the COVID-19 outbreak and throughout the pandemic, the present study aims to examine change in MVC patterns and associated injuries and deaths in 2019 and 2020. This research specifically focuses on speeding-related MVCs, while comparing this crash type to other types of MVCs (i.e., collisions involving pedestrians and cyclists and single-vehicle collisions) that are often associated with more severe and fatal injuries.^12–14^ By controlling for the reduction of traffic volumes and seasonality, we sought to understand if the risk of experiencing a traffic-related injury or fatality would be lower during the pandemic, and how these changes would vary from the first to the second phase of the pandemic when some of the restrictions were lifted.

## DATA AND METHODS

We collected traffic collision datasets from NYC Open Data and City of Seattle Open Data programs.^15,16^ Both datasets contain information regarding MVCs which occurred between January 1, 2019, and December 31, 2020, submitted by law enforcement. There was a total of 321,816 MVCs in NYC and a total of 16,446 MVCs in Seattle during the study period. The collision data include the number of motorists, passengers, pedestrians, and cyclists involved; major contributing factors (e.g., speeding); severity (i.e., property damage only, injury, severe injury, or death); and others (e.g., location and time). The information about speeding was explicitly included in the database maintained by Seattle Police Department and “unsafe speed” was considered as one of the major contributing factors in the dataset originally from New York Police Department. For the following difference-in-differences (DID) analysis considered in the study, we aggregated the raw crash data into a weekly number of collisions, people injured and killed, or severely injured (only available in the dataset from Seattle). According to the data source, the severe injury was defined as a “Serious bodily injury… which causes a temporary but substantial loss or impairment of the function of any bodily part or organ.”

This study employed a DID econometric approach to quantify whether weekly trends of MVCs differed significantly from those of 2019 when COVID19 did not occur. The empirical model assumes that in the absence of the pandemic, temporal changes in MVCs before and after the onset of the COVID-19 outbreak would be essentially the same in 2020. Therefore, the year 2020 is considered as the “treatment group” and the periods after the disruption in both 2019 and 2020 are considered as the “aftertreatment periods.” ^4,17^ A similar quasi-experimental approach has been employed in previous studies focusing on various impacts of the pandemic (e.g., air quality).^4^ The DID based Poisson regression is specified by the following equation:

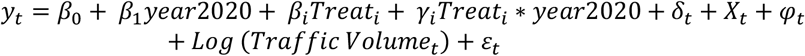

where the dependent variable *y*_*t*_ is the MVC outcomes in weekly counts; year2020 is a dummy variable indicating the year of 2020 (coded=1); *Treat*_*i*_ equals the ‘treatment periods’ that take the value of 1 for calendar months from March to May and the value of 2 for calendar months from June to December. *γ*_*i*_equals the interactions between the treatment group and the aftertreatment-period term, which is the main variable of interest. The two “aftertreatment-periods” were specified based on the timeline of the two phases of the pandemic,^18^ while following the results of change-point detection using daily traffic volume data (online supplementary Figure 1). *δ*_*t*_ equals the trend term; *φ*_*t*_ equals the monthly dummies; *X*_*t*_ refers to covariates of average daily precipitation and average daily maximum temperature included to control for seasonal weather conditions.^5,9^

We included the log of weekly traffic counts as a proxy of exposure. Traffic flow data in Seattle was retrieved from loop detectors at major highway entrances of Interstates I-5 and I-90, which measure commuter traffic to or from the city of Seattle. Likewise, we gathered the dataset containing the hourly volume of all vehicles passing through each of the nine bridges and tunnels in NYC operated by the Metropolitan Transportation Authority (MTA).^19^

## RESULTS

Figures 1 and 2 show weekly trends of traffic volumes and MVC outcomes in New York City and Seattle in 2019 and 2020. There appeared to be steep declines in total crashes and traffic volumes during the period of March-May or the period during relatively stringent stay-at-home restrictions. The rebound of vehicle volumes might have started as early as April, while they remained 20-30% lower compared to the level of 2019. Largely due to the reduction in traffic volume, Seattle saw a 38.3% decrease in traffic-related injuries or 1,608 fewer injuries in 2020; in New York City, there was a 28.0% decline or 17,108 fewer traffic-related injuries during the same period. It should be noted that the number of fatal or severe injury crashes only declined substantially during the first phase of the pandemic. In the second half of 2020, or after both cities lifted some of the restrictions, there were a 12.71% reduction of traffic-related severe injuries or fatalities in Seattle and a 27.65% increase of fatal MVCs in NYC, compared with the decreases of 39.2% (Seattle) and 49.21% (NYC) in all type crashes (online supplementary Tables 1 and 2). In fact, the death toll associated with MVCs in NYC increased from 244 in 2019 to 265 in 2020, and the number of traffic-related fatalities remained largely unchanged in Seattle (26 in 2019 and 24 in 2020).

**Figure 1.**
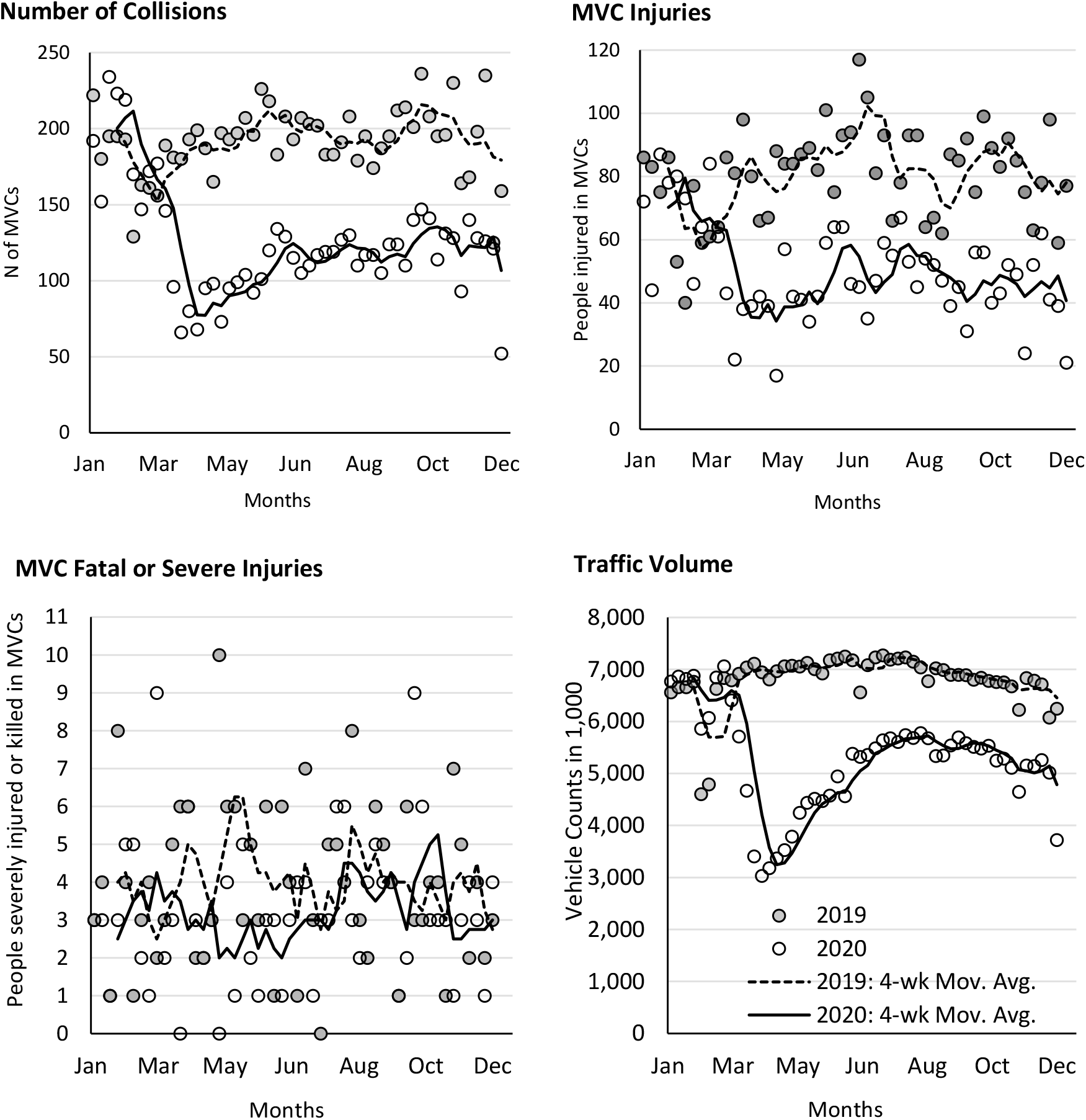
Weekly Counts of Vehicle Crash-Related Outcomes and Traffic Volume (highway loop detectors) in Seattle, WA during 2020 COVID-19 Pandemic.

**Figure 2.**
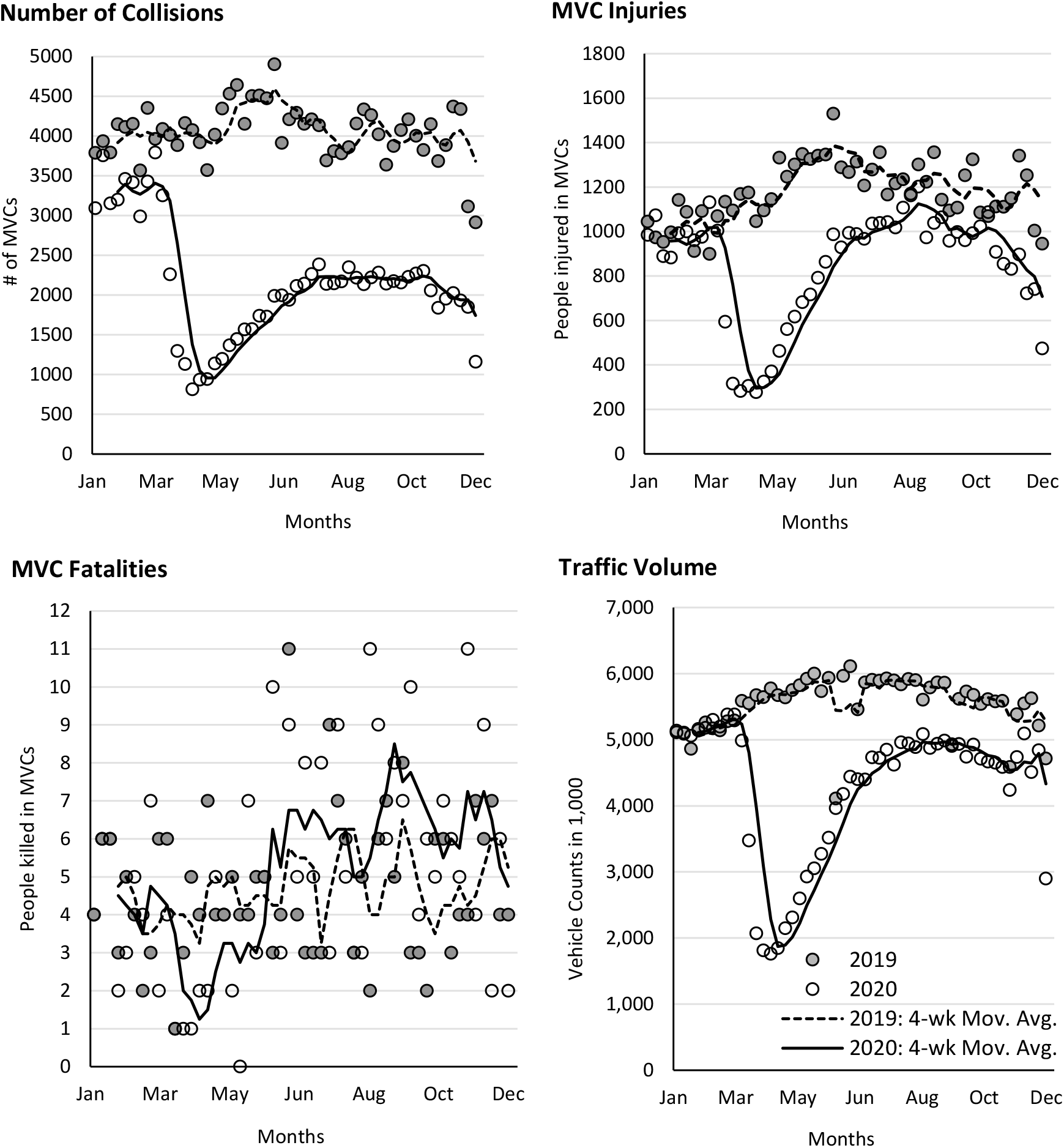
Weekly Counts of Vehicle Crash-Related Outcomes and Traffic Volume (nine tunnels and bridges) in New York City, NY during 2020 COVID-19 Pandemic.

Sixteen DID regression models were computed using STATA software, and the results are summarized in Table 1. Several interesting findings emerge: First, after controlling for reductions in traffic flows and seasonal weather conditions, total crash rates were significantly lower than those of 2019 during the second phase of the pandemic, but fatal crash rates in NYC significantly increased 1.468 times (95% CI 1.044 to 2.065) in the second phase. Second, the expected incidence rates of speeding-related collisions significantly increased. The crash rates of speeding-related collisions that cause fatal or severe injuries increased 7.7 times in Seattle (95% CI 1.321 to 44.416) and 4.5 times in NYC (95% CI 1.196 to 17.108) during the months of March-May. In the second phase of the pandemic (i.e., June-December), they persisted higher than what would be expected in the absence of the pandemic (NYC: IRR 2.187, 95% CI 1.070 to 4.467; Seattle: IRR 4.158, 95% CI 1.598 to 10.818). Third, no significant changes were found for the fatal or severe injury crash rates of collisions that involve pedestrians or cyclists, or single-vehicle collisions throughout the pandemic in 2020.

**Table 1.** Results of Difference-in-Differences (DID) Regressions for Crashes in Seattle and New York City during 2020 COVID-19 Outbreak.

| Outcome models | Treatment period of Mar.-May (DID interaction terms, IRR) |  |  |  | Treatment period of Jun.-Dec. (DID interaction terms, IRR) |  |  |  |
| --- | --- | --- | --- | --- | --- | --- | --- | --- |
|  | (1) | (2) | (3) | (4) | (1) | (2) | (3) | (4) |
|  | All type | Speeding | Single vehicle | Ped. & Cyclists | All type | Speeding | Single vehicle | Ped. & Cyclists |
| **Seattle** |  |  |  |  |  |  |  |  |
| # of crashes | 0.923<br>(0.821 to 1.038) | 3.349***<br>(1.566 to 7.163) | 1.137<br>(0.967 to 1.336) | 0.689*<br>(0.467 to 1.016) | 0.848***<br>(0.800 to 0.899) | 1.263<br>(0.647 to 2.468) | 0.934<br>(0.828 to 1.053) | 0.668*<br>(0.445 to 1.002) |
| Injuries | 0.976<br>(0.759 to 1.255) | 2.635***<br>(2.000 to 3.472) | 1.056<br>(0.799 to 1.395) | 0.665*<br>(0.437 to 1.009) | 0.833<br>(0.661 to 1.026) | 1.161<br>(0.497 to 2.715) | 0.763**<br>(0.583 to 0.997) | 0.657**<br>(0.451 to 0.955) |
| Fatal/severe injuries | 1.354<br>(0.655 to 2.797) | 7.661**<br>(1.321 to 44.416) | 1.944*<br>(0.972 to 3.890) | 0.848<br>(0.308 to 2.335) | 1.452*<br>(0.944 to 2.233) | 4.158***<br>(1.598 to 10.818) | 1.705<br>(0.778 to 3.605) | 1.166<br>(0.612 to 2.223) |
| **New York City** |  |  |  |  |  |  |  |  |
| # of crashes | 0.922<br>(0.750 to 1.133) | 1.720***<br>(1.379 to 2.145) | 1.285***<br>(1.139 to 1.449) | 1.094<br>(0.932 to 1.212) | 0.745***<br>(0.696 to 0.797) | 1.276***<br>(1.168 to 1.395) | 1.003<br>(0.882 to 1.141) | 1.126*<br>(0.998 to 1.269) |
| Injuries | 0.993<br>(0.901 to 1.052) | 1.705***<br>(1.263 to 2.303) | 1.034<br>(0.969 to 1.103) | 1.061<br>(0.925 to 1.236) | 1.022<br>(0.949 to 1.101) | 1.730***<br>(1.438 to 2.081) | 0.905<br>(0.788 to 1.039) | 1.135*<br>(0.995 to 1.296) |
| Fatal injuries | 1.202<br>(0.718 to 2.011) | 4.523**<br>(1.196 to 17.108) | 0.619<br>(0.226 to 1.695) | 0.898<br>(0.553 to 1.457) | 1.468**<br>(1.044 to 2.065) | 2.187**<br>(1.070 to 4.467) | 1.023<br>(0.553 to 1.894) | 1.269<br>(0.923 to 1.745) |

Note: Model specified a Poisson distribution of the outcome, and a log transformation of traffic flow was included as an exposure variable. Clustered SEs were used for a more conservative estimate of the IRR or incidence rate ratio. Average daily precipitation and average daily highest recorded temperatures, linear time, and monthly dummies were included in the regressions. Climate data were retrieved from the National Climate Data Center's climate data online database, and we collapsed recorded daily maximum temperature and average precipitation into weekly averages from weather stations in New York City and Seattle.
95% CIs are reported in brackets. Ped, pedestrians; IRR, incidence rate ratio. \*\*\*p<0.01, \*\*p<0.05, \*p<0.1

## DISCUSSION

Despite the relative reduction of traffic-related collisions and injuries, the death toll associated with MVCs in 2020 remained largely unchanged in New York City and Seattle. Speeding-related crashes that cause severe or fatal injuries were found to increase substantially. This result is largely attributed to the decreased traffic volume because of the lockdown and teleworking, which would have created an encouraging environment for speeding. However, other factors like alcohol consumption, depression, and suicidality might also prompt high-risk driving behavior during the pandemic, which warrants further investigation.^20^

One of the advantages of our study is that we employed a DID econometric approach to model crash occurrence, while accounting for exposure by using a traffic volume proxy that exhibited reductions during the pandemic. Moreover, instead of analyzing records on traffic violations or speeding tickets, speeding is directly measured as a contributing factor of MVCs by using crash data.^10^

Our results suggest that when traffic demand abruptly decreases due to emergencies like natural disasters, pandemics, or business shutdowns, speeding behavior will be more prevalent. Under these circumstances, efforts to promote road safety should consider countermeasures specifically targeting speeding violations. Policy makers might consider installing temporary traffic calming devices near high-risk locations, increasing speed enforcement, installing speed trailers, and employing rapid response education campaigns and public service announcements. These measures may help road users to avoid speeding and in turn reduce severe injuries and fatalities.

### Bullet points

#### What is already known on the subject

- There has been a decrease in traffic collisions and related injuries during the COVID-19 pandemic.
- Low traffic volume during this period would prompt drivers to speed on empty roads and cause traffic-related fatalities and severe injuries.

#### What this study adds

- After controlling for reductions in traffic volume, this study estimates crash rates stratified by injury severity during the 2020 COVID-19 pandemic, with a particular focus on speeding-related collisions.
- Despite a relative decrease in MVCs and injuries during the early months of the outbreak, fatal or severe injury crash rates of speeding related MVCs increased 7.7 times (95% CI: 1.321 to 44.416) in Seattle, and 4.5 times (95% CI: 1.196 to 17.108) in NYC, compared with what would be expected in the absence of the pandemic.
- During the second phase of the pandemic when some of the restrictions were lifted, the increases of speeding-related collisions remained significantly higher than expected, in conjunction with significant increases in fatal crash rates in NYC (IRR 1.468; 95% CI 1.044 to 2.065).

## Supporting information

Changes in Crashes and Traffic-Related Injuries or Fatalities in Seattle and NYC during 2020 COVID-19 Outbreak

Supplementary Figure 1. Top: Breaking Points of Traffic Flows in Seattle (major highway locations) Bottom: Breaking Points of Traffic Flows in New Yor

## Data Availability

not applicable

## Statements

## Acknowledgments

This study was funded by a grant from Pacific Northwest Transportation Consortium (Pac Trans), USDOT Transportation Center for Federal Region 10 under the award #69A3551747110.

## Ethics statements

### Patient consent for publication

Not required.

### Ethics approval

The study used publicly available and deidentified secondary data reported on an aggregated level. Therefore, ethics approval was not required.

## Contributions

FL conceived and designed the study, obtained and analyzed the data, and drafted the manuscript. ML made comments for revisions of the manuscript, and all authors approved of the current version.

