## Supplementary material for "Speeding and Traffic-Related Injuries and Fatalities during the 2020 COVID-19 Pandemic: The Cases of Seattle and New York City": Changes in Crashes and Traffic-Related Injuries or Fatalities in Seattle and NYC during 2020 COVID-19 Outbreak

**Supplementary Table 1. Changes in Crashes and Traffic-Related Injuries or Fatalities in Seattle, WA during 2020 COVID-19 Outbreak**

| | Jan-Feb | | March-May | | | June-December | | | |
|---|---|---|---|---|---|---|---|---|---|
| | Mean Weekly Counts | vs the same period in 2019 | Mean Weekly Counts | vs the same period in 2019 | vs Jan-Feb | Mean Weekly Counts | vs the same period in 2019 | vs Jan-Feb | vs March-May |
| **# of crashes** | 188.63 | 4.94% | 99.15 | -47.17% | -47.43% | 119.16 | -39.20% | -36.83% | 20.18% |
| Speeding | 7.75 | 3.33% | 7.85 | 70.00% | 1.24% | 5.97 | -22.28% | -23.01% | -23.95% |
| Single vehicle | 27.50 | -5.58% | 16.08 | -42.11% | -41.54% | 19.19 | -39.41% | -30.21% | 19.39% |
| Ped. or cyclists | 14.63 | -4.88% | 6.15 | -65.37% | -57.92% | 8.23 | -55.73% | -43.76% | 33.67% |
| **# of injuries** | 68.00 | -2.68% | 42.86 | -46.17% | -36.97% | 47.87 | -42.95% | -29.60% | 11.70% |
| Speeding | 3.88 | -3.13% | 4.15 | 25.58% | 7.20% | 3.19 | -35.29% | -17.59% | -23.12% |
| Single vehicle | 16.88 | -4.26% | 9.62 | -50.00% | -43.02% | 10.35 | -49.77% | -38.64% | 7.69% |
| Ped. or cyclists | 13.00 | -5.45% | 5.54 | -67.27% | -57.40% | 7.32 | -57.57% | -43.67% | 32.21% |
| **# of severe or fatal injuries** | 2.88 | -17.86% | 3.08 | -32.20% | 7.02% | 3.32 | -12.71% | 15.57% | 7.98% |
| Speeding | 0.75 | -25.00% | 0.85 | 175.00% | 12.82% | 0.65 | 81.82% | -13.98% | -23.75% |
| Single vehicle | 1.50 | -33.33% | 1.77 | -30.30% | 17.95% | 1.94 | -18.92% | 29.03% | 9.40% |
| Ped. or cyclists | 1.25 | -33.33% | 0.92 | -65.71% | -26.15% | 1.39 | -35.82% | 10.97% | 50.27% |
| **Traffic flow (in million)** | 6.646 | 7.42% | 4.238 | -39.34% | -36.23% | 5.316 | -22.91% | -20.01% | 25.43% |

Ped, pedestrians.

**Supplementary Table 2. Changes in Crashes and Traffic related Injuries or Fatalities in New York City during 2020 COVID-19 outbreak**

| | Jan-Feb | | March-May | | | June-December | | | |
|---|---|---|---|---|---|---|---|---|---|
| | Mean Weekly Counts | vs the same period in 2020 | Mean Weekly Counts | vs the same period in 2020 | vs Jan-Feb | Mean Weekly Counts | vs the same period in 2020 | vs Jan-Feb | vs March-May |
| **# of crashes** | 3312.38 | -16.82% | 1623.43 | -60.46% | -50.99% | 2053.23 | -49.21% | -38.01% | 26.47% |
| Speeding | 67.25 | 2.48% | 61.43 | -9.36% | -8.66% | 75.29 | 12.10% | 11.96% | 22.57% |
| Single vehicle | 298.88 | -16.46% | 170.00 | -44.61% | -43.12% | 230.70 | -93.76% | -22.81% | 35.71% |
| Ped/cyclists | 287.13 | -4.89% | 153.08 | -50.03% | -46.69% | 296.03 | -14.74% | 3.10% | 93.39% |
| **Injuries** | 970.13 | -5.42% | 533.46 | -53.96% | -45.01% | 948.00 | -22.51% | -2.28% | 77.71% |
| Speeding | 39.25 | -6.55% | 37.15 | -26.71% | -5.34% | 58.10 | 30.70% | 48.02% | 56.37% |
| Single vehicle | 225.00 | -9.04% | 104.00 | -53.06% | -53.78% | 163.52 | -31.86% | -27.33% | 57.23% |
| Ped/cyclists | 263.00 | -5.40% | 137.00 | -51.25% | -47.91% | 274.81 | -14.25% | 4.49% | 100.59% |
| **Fatal injuries** | 4.63 | 12.12% | 2.62 | -41.38% | -43.45% | 6.30 | 27.65% | 36.22% | 140.88% |
| Speeding | 1.00 | 14.29% | 1.08 | 75.00% | 7.69% | 1.33 | 65.33% | 33.33% | 23.81% |
| Single vehicle | 2.50 | -4.76% | 0.79 | -70.82% | -68.57% | 2.37 | -14.69% | -5.33% | 201.21% |
| Ped/cyclists | 2.75 | -4.35% | 1.00 | -63.89% | -63.64% | 3.13 | -5.83% | 13.78% | 212.90% |
| **Traffic flow (in millions)** | 5.187 | 1.04% | 2.940 | -48.39% | -43.32% | 4.648 | -16.79% | -10.39% | 58.10% |

Ped, pedestrians.
