## Supplementary Figure 1. Top: Breaking Points of Traffic Flows in Seattle (major highway locations) Bottom: Breaking Points of Traffic Flows in New Yor for "Speeding and Traffic-Related Injuries and Fatalities during the 2020 COVID-19 Pandemic: The Cases of Seattle and New York City"

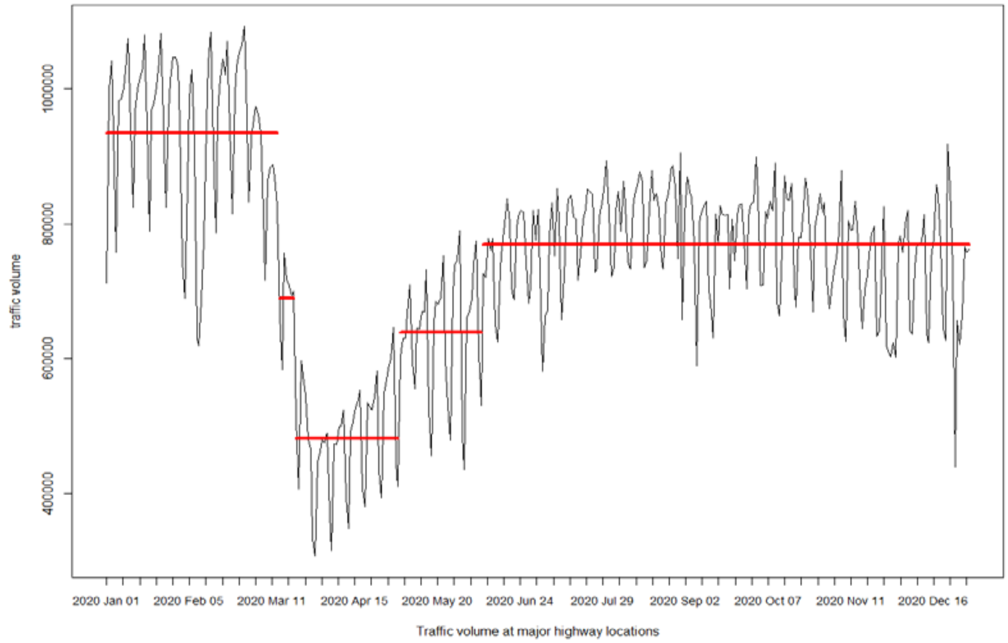


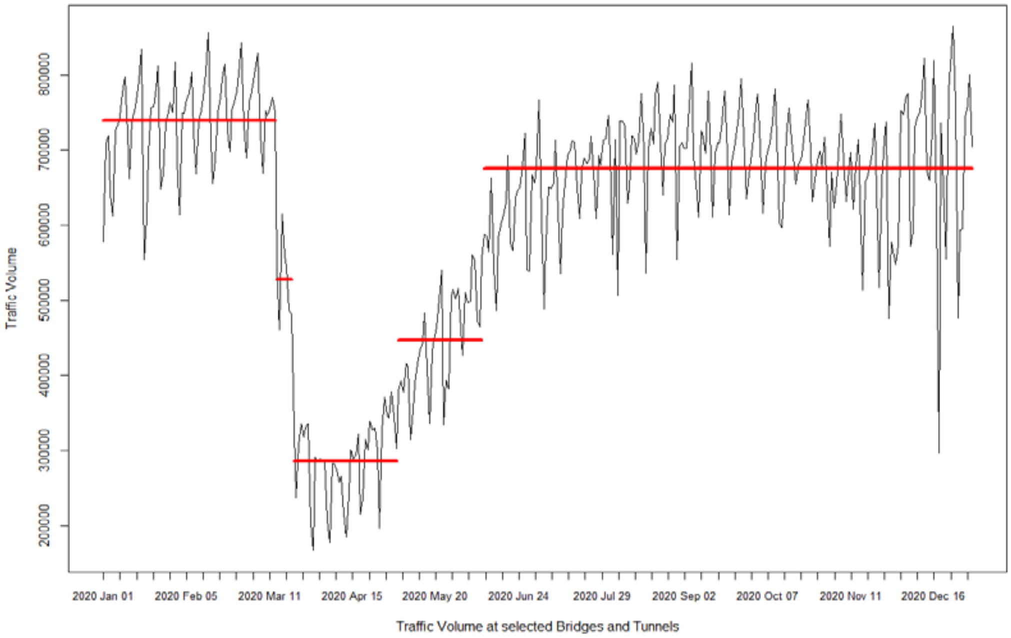


**Supplementary Figure 1. Top: Breaking Points of Traffic Flows in Seattle (major highway locations) Bottom: Breaking Points of Traffic Flows in New York City (nine bridges and tunnels)**

Note: We applied the R package 'strucchange', which offers a collection of tools for detecting changes within linear regression models. The breaking point estimation and the Bayesian Information Criteria-based optimization function were used for determining the number and time of change points.

Red line: BIC Regression Breaking Point.
